# Leveraging hierarchical structures for genetic block interaction studies using the hierarchical transformer

**DOI:** 10.1101/2024.11.18.24317486

**Authors:** Shiying Li, Shivam Arora, Redha Attaoua, Pavel Hamet, Johanne Tremblay, Alexander Bihlo, Bang Liu, Guy Rutter

**Affiliations:** Centre de Recherche du CHUM, and Faculty of Medicine, University of Montreal, QC, Canada; Department of Mathematics and Statistics, Memorial University of Newfoundland, NL, Canada; Département d’informatique et de recherche opérationnelle, Université de Montréal, QC, Canada; Section of Cell Biology and Functional Genomics, Department of Metabolism, Diabetes and Reproduction, Imperial College of London, du Cane Road, London W120NN, United Kingdom; Lee Kong Chian School of Medicine, Nan Yang Technological University, Singapore

## Abstract

Initially introduced in 1909 by William Bateson, classic epistasis (genetic variant interaction) refers to the phenomenon that one variant prevents another variant from a different locus from manifesting its effects. The potential effects of genetic variant interactions on complex diseases have been recognized for the past decades. Moreover, It has been studied and demonstrated that leveraging the combined SNP effects within the genetic block can significantly increase calculation power, reducing background noise, ultimately leading to novel epistasis discovery that the single SNP statistical epistasis study might overlook. However, it is still an open question how we can best combine gene structure representation modelling and interaction learning into an end-to-end model for gene interaction searching. Here, in the current study, we developed a neural genetic block interaction searching model that can effectively process large SNP chip inputs and output the potential genetic block interaction heatmap. Our model augments a previously published hierarchical transformer architecture (Liu and Lapata, 2019) with the ability to model genetic blocks. The cross-block relationship mapping was achieved via a hierarchical attention mechanism which allows the sharing of information regarding specific phenotypes, as opposed to simple unsupervised dimensionality reduction methods e.g. PCA. Results on both simulation and UK Biobank studies show our model brings substantial improvements compared to traditional exhaustive searching and neural network methods.

## 2. Introduction

In the past decades, the genetic factors that contribute to the pathogenesis of complex diseases have been extensively studied, and genome-wide association study (GWAS) has played a fundamental role in unveiling novel risk alleles. The approach of the GWAS framework assumes each SNP has an independent effect on phenotype and the disease’s statistical relevance was tested individually (Niel et al., 2015). However, most complex disease variants identified so far confer relatively small increments in risk, leading to many questions about how the remaining “missing heritability” can be explained (Maher, 2008). For example, GWAS correlation identified >80 common variants for type 2 diabetes with most of those associated with insulin secretion, together, they only contribute ∼ 10% of type 2 diabetes (T2D) heritability. T2D Low-frequency and rare variants have also been identified, but their contribution towards “missing heritability” is also limited (Stančáková and Laakso, 2016). Indeed, with a larger sample size and the advancement of sequencing techniques, GWAS will likely continue to expand the number of novel complex disease genetic markers. However, the current consensus underscores the growing recognition that the missing heritability of complex diseases extends beyond the scope of singular genetic factors. Interactions among two or more SNPs, a combinatorial effect known as epistasis, have been proven (Turton et al., 2011) can at least partly explain the “missing heritability”.

Finding the optimal interacting SNP combination for certain phenotypes, which implies an exhaustive search of all possible cases, can be a challenging task. For instance, in a dataset containing 500,000 SNPs, there are approximately 250 billion possible pairwise SNP combinations. This immense number presents significant challenges, not only in terms of computational hardware requirements but also in the risk of losing true signals due to overcorrection for multiple comparisons resulting in a reduction in statistical power. A problem we often refer to as the “curse of dimensionality”. To address this issue, several scalable statistical approaches have been proposed in recent years; however, each comes with its own set of limitations. Some methods (Cordell, 2002) only select “top SNPs” (the SNPs most correlated with phenotypes) for epistasis searching, while ignoring the potential effects of neighbouring SNPs. Indeed, over the past few decades, extensive research (Morris and Kaplan, 2002; Zaykin et al., 2002; Chen et al., 2020) has highlighted the necessity and efficiency of leveraging the combined effects of multiple SNPs within certain genetic regions e.g. haplotype blocks, rather than focusing on individual SNPs in association studies. Some have proposed to summarise multi-dimensional SNPs into one-dimensional representations using unsupervised methods such as PCA (Li et al., 2009). However, these methods overlook important phenotype information and compress highly dependent SNPs into a single dimension, making it difficult to detect signals within these units. In short, it is still an open question how we can best combine genetic block representation learning and interaction modelling to an end-to-end model to increase calculation power.

The rapid development of deep learning and artificial intelligence seems to hold another promise for epistasis studies. Several studies (Pérez-Enciso and Zingaretti 2019; Cui et al., 2022) have suggested Deep Neural Networks (DNNs) can map the flexible, both linear and non-linear relationships between SNPs and observed phenotypes. However, DNNs, which are primarily designed for classification, with a black-box nature that makes it challenging to interpret results, particularly in identifying which SNPs are interacting. Many studies aim to bridge this gap between interpretability and DNN structures using such as layerwise relevance (Mieth et al., 2021), permutation testing (Cui et al., 2022), and more recently, transformer with attention scores (Graça et al., 2024). While the potential application of transformer in genome sequencing analysis has enjoyed renewed interest. Scanning through most of the genetic transformer studies in recent years (Jubair et al., 2021; Reyes et al., 2022; Zhou et al., 2022), the basic unit, as the “word” in natural language, is still single SNP with little existing genetic structure e.g. haplotype block, the main focus of current study, introduced within the attention block. The size of haplotype blocks can vary and is often larger than the typical units analysed in natural language processing. This variability poses a substantial challenge in encoding these differently-sized haplotype blocks into the transformer encoder while preserving their biological significance during this process, to ensure that the outputs of the attention scores are interpretable and relevant.

In the current study, taking inspiration from the hierarchical transformer model (Liu and Lapata, 2019), we proposed a novel haplotype block-haplotype block association study workflow, Haplotype Block LSTM hierarchical Transformer (HB-LT). HB-LT is constructed in a hierarchical manner which allows it to efficiently capture both within and cross-haplotype relationships relevant to specific phenotypes. We demonstrate with simulations that grouping SNPs into dimensionality-reduced haplotype block structures significantly increases detection power for epistasis studies compared to existing methods. Furthermore, by evaluating our model on the UK Biobank dataset, we demonstrated its potential for real-world applications.

## 3. Methods

### 3.1 Model description

The model in the current study is mainly inspired by Liu and Lapata (2019) and several previous machine learning works (Chang et al., 2020; Cui et al., 2022 and Graça et al., 2023) dedicated to epistasis studies. Our haplotype epistasis study system is illustrated in Figure 1. The inputs of the model are the pre-organised haplotype datasets and the associated phenotype of each individual, while the outputs are the attention weights (epistasis) among potential candidates.

**Figure 1.**
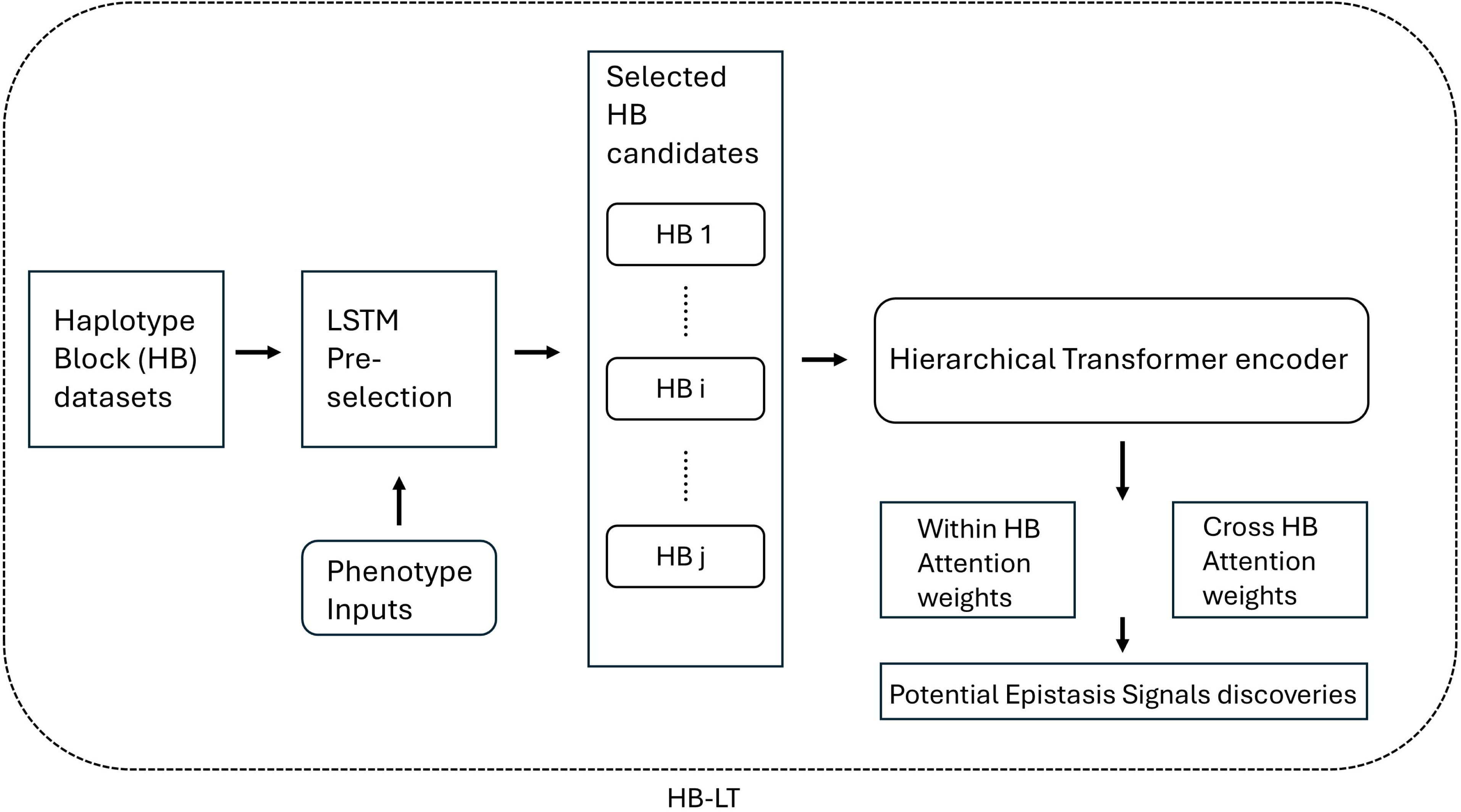
Pipeline of our within and cross haplotype block epistasis detection method (HB-LT). The haplotype dataset was first selected by Long short-term memory (LSTM) to obtain the potential phenotype-associated haplotype blocks (HBs). These candidates are then fed into the Hierarchical transformer encoder to obtain the within and cross-haplotype block attention weights, which could be subjected to the following analysis for potential epistasis signal discoveries.

#### 3.1.1 Long short-term memory (LSTM) pre-selection

For large dataset analysis, applying pre-selection methods effectively reduces computational burdens and enhances calculation efficiency. However, one of the key challenges in the current study is the variation in haplotype block lengths, which can range from as few as 2 to more than 100 SNPs. Here, we adopted a learning-based approach. A linear regression model is applied to each haplotype individually, and its average root of mean square error on the testing dataset is used as a score indicating whether it should be selected as a phenotype-associated candidate. Haplotype blocks of the SNP dataset were first constructed using the confidence interval method (Gabriel et al., 2002), we then use recurrent neural network LSTM to represent each haplotype block. Let

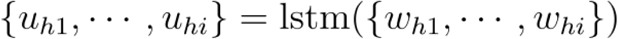

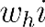 are word embedding for tokens in each haplotype block, where 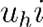 are updated vectors for the token after LSTM.

An average-pooling is then used to obtain a fixed length representation and a linear transformation yields the final representation of the haplotype block *s_i_*.

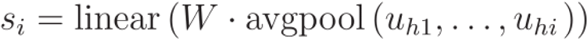

All input haplotype blocks were pre-split into training and testing datasets. The model is trained by minimising the root of mean square error of *s_i_* and the phenotype *y*. In testing, the phenotype-associated haplotype block candidates were selected based on the mean prediction score.

#### 3.1.2 Hierarchical transformer encoder

##### 3.1.2.1 Embedding

Input SNPs are first represented by word embeddings. Let 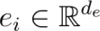 represent the embedded dimensional vectors of the SNP *i*. Let *H* denote the haplotype where 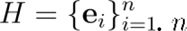 is the total number of SNPs in each haplotype block. In our hierarchical haplotype transformer, each token (SNP) has two positions that need to be considered, namely *i*, the position of the token (SNP) within the haplotype, and *j*, the position of the haplotype block within the input sequence. We follow (Vaswani et al., 2017) and use sine and cosine functions for calculating positional embedding. These two positional embedding vectors were then concatenated and the final input vector of each token (SNP) for the hierarchical haplotype transformer model is: 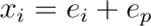.

##### 3.1.2.2 Local haplotype attention block

The main aim of the local haplotype attention block is to map the dynamic attention scores among SNPs within each haplotype block. It contains several components, including multi-head attention, layer normalisation and feed-forward. The number of these local attention blocks to be used in the model will be decided by the researchers themselves. Let 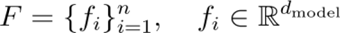 denote the features of SNPs within each haplotype input to the local haplotype attention block. For the *i* attention head, the query 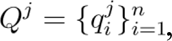, key 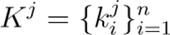 and value 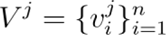 of each SNP are calculated based on 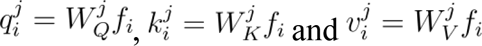 respectively. The linear projection learnable parameters weight *W* are matrices 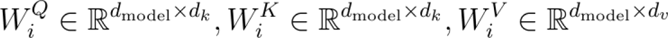. 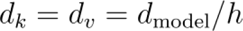. The output of *j* attention head will be: 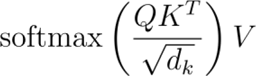. The multi-head results were then concatenated and linear transferred with learnable weight 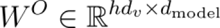 to get the final results. The feed-forward layer is composed of two fully connected (FC) layers in inverse order with an activation function *Tahn* in between.

##### 3.1.2.3 Inter-haplotype attention block

To exchange information across different haplotypes, an inter-haplotype attention block was used. To obtain a fixed-length haplotype representation, a weighted, multi-head pooling was first used to represent each haplotype. In each head, weight distributions over tokens (SNPs) are calculated and different heads will encode haplotypes with different attention weights. Let 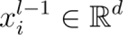 denote the output vector from the last layer of the local haplotype attention block, which will be the input of the multi-head pooling layer. For haplotype *H_j_*, for head *z*, a linear transformation was first applied to convert the input vector into an attention score 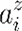 and a value vector 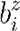 with the weight 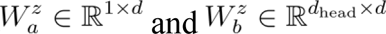. The final output *d*_head_ x 1 is a weighted sum representing haplotype *j* in head *z* where 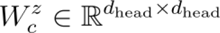.

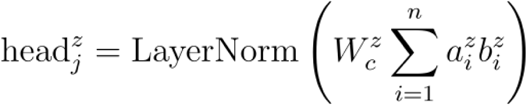

Similar to local haplotype attention blocks, inter-haplotype allows one haplotype to attend to another to model the haplotype-haplotype dependencies.

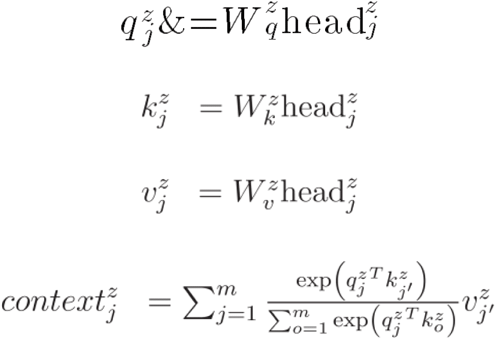

Where 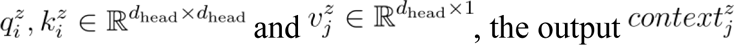, was then flatted to generate a vector with dimension 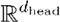. Finally, the different heads for each haplotype were then concatenated and linear transformed 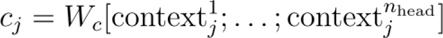 where 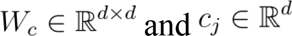 will be added to the original token *i* vector to update the token. Figure 2 provides a schematic view of inter-haplotype block attention.

**Figure 2.**
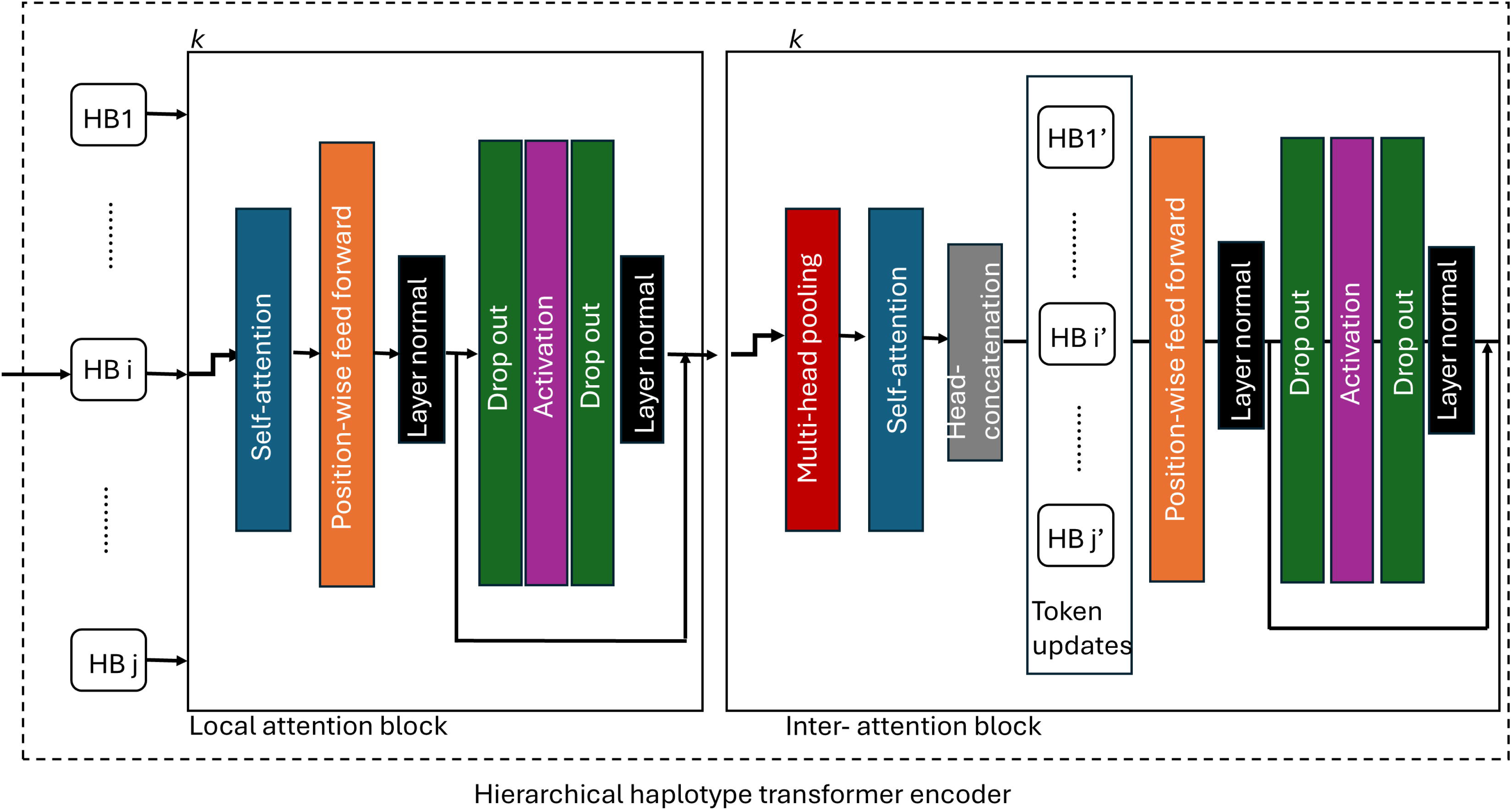
The overall pipeline of the hierarchical haplotype transformer encoder.

### 3.2 Simulation dataset

The simulation datasets were achieved by re-sampling approaches with existing genotype data as reference panels, thereby retaining allele frequency and LD patterns (Wright et al., 2007). In the current study, the re-sampling based method Hapgen2 (Su et al., 2011) was applied with 1000Genomes (Auton & Salcedo, 2015) as a reference panel. In total, chromosome 20 of 1000 individuals was resampled and subjected to the following analysis. The haplotype block was parsed based on the confidence interval method (Gabriel et al., 2002) by *PLINK* (Purcell et al., 2007).

In our current study, we first simulate the expression of *h_i_* of each haplotype block according to a linear combination of all SNPs in the haplotype block.

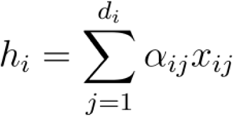

The phenotype was then simulated based on three epistasis models, which were originally proposed by Burton et al (2007). More specifically, model 1 reflects an epistasis model where the odds of disease increase multiplicatively within and between 2-way disease markers. Using *h_i_* and *h_j_* to denote the expression of haplotype and *_i_*, and *j, α* and *θ* to denote the baseline and the factor of odd disease increase. Model1:

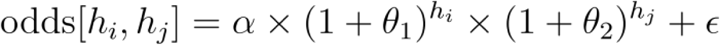

In contrast, model 2 represents a disease model where the odds of disease only increase unless both loci have at least one disease-associated allele,

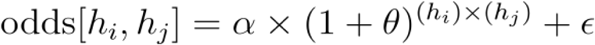

Model 3 is similar to model 2, but renders a simpler threshold model as

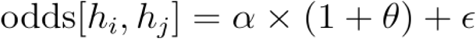

To simulate the simple epistasis model, two haplotype blocks will be randomly selected each time and the phenotype model 1, model 2 or model 3 will be simulated accordingly. To simulate the complex epistasis model, one dataset will contain multiple epistasis from different SNP pairs. Here, we use the ‘Combined Model 1+2+3’ as a complex epistasis model which contains three epistasis from the previous three basic models.

### 3.3 Baseline models

To benchmark our model, we selected two state-of-the-art approaches for comparison with the current proposed framework, matrixEpistasis (Zhu and Fang, 2017) and GWAS_NN (Cui et al., 2022). MatrixEpistasis represents a state-of-the-art method for exhaustive epistasis searching. In contrast, GWAS_NN is one of the few methods in the current field that tackles epistasis detection using neural networks, while also providing an interpretation for the observed results. The GWAS_NN model first learns the genetic block representations from all SNPs of a genetic block in a shallow layer and then learns the complex relationships between genetic blocks in a deep layer. These two baseline models exemplify the two main categories for epistasis detection: exhaustive searching and machine learning. Both baseline models were operated with the default settings unless indicated otherwise.

### 3.4 Cohort description and statistical analysis

#### 3.4.1 Cohort description

This research has been conducted using the UK Biobank Resource. A material transfer agreement was signed with UK Biobank that covers Research Tissue Bank (RTB) under projects 49731 and 59642. The UK Biobank study began in 2006 and, by 2010, had recruited over 500,000 participants from the general UK population, aged 40 to 69 at the time of enrollment. The UK Biobank genetic data contains genotypes for 488,377 participants. These were assayed using two different yet similar assays, Applied Biosystems UK BiLEVE Axiom Array by Affymetrix (Thermo Fisher Scientific) and Applied Biosystems UK Biobank Axiom Array. More detail on the assay and quality control can be found in the UK Biobank Genotyping and Quality Control (https://biobank.ctsu.ox.ac.uk/crystal/crystal/docs/genotyping_qc.pdf.). Individuals included in the current study from UK Biobank have T2D and are of European descent. The UK The REC reference for UK Biobank is 16/NW/0274.

#### 3.4.2 Candidate genes selection

To test the potential application of HB-LT in a real-world scenario, we applied HB-LT to pre-selected glycated haemoglobin associated genes. The candidate genes were first extracted from the DisGeNET database (Piñero, et al., 2015). In total, fourteen genes were extracted and their coordinates (chromosome, gene start position, and gene end position) were obtained using the BioMart Project martview tool (Supplemental material). Next, SNPs located in each gene ± 10 kbps were extracted in *PLINK* (Purcell et al., 2007). The thresholds set for quality control including, imputation quality, Hardy-Weinberg equilibrium, genotyping missing data across individuals, and genotyping missing rate were 0.8, 10-10, 0.05, and 0.05 respectively.

#### 3.4.3 Covariants pre-filtering

The datasets were subjected to a PCA-based covariant pre-filtering stage to reduce the confounding effects before they feed into the HB-LT for potential epistasis signal mining. Four covariants, including sex, age of diabetes diagnosis, diabetes duration and population genetic structure were standardised (*Scikit-learn* package) and subjected to PCA (*Scikit-learn* package) to reduce into a 2-dimensional space. A× 1 square was then applied to locate the most densely populated area and individuals within this area were selected and subjected to the following analysis.

### 3.5 Software support

We conducted model-building and statistical analysis mainly using Python 3.9 (https://www.python.org/) and additional packages including *Pandas* (2.2.2), *Numpy* (1.26.4), *PyTorch* (2.3.1), and *Tensorflow* (2.16.1). Other software includes R 4.2.2 (https://www.r-project.org/), Hapgen2 (https://mathgen.stats.ox.ac.uk/genetics_software/hapgen/), PLINK 1.9 (https://www.cog-genomics.org/plink/) and BioMart Project martview (https://mart.ensembl.org/). The figures in the current study were drawn by *Matplotlib* (3.8.2) and *Plotly* (5.22.0).

### 3.6 Data availability statement

UK Biobank data are available to registered investigators under approved applications (http://www.ukbiobank.ac.uk). Other relevant data are available from the corresponding author upon request. The source code is available at https://github.com/LI-SHIYING118/HB-LT.

## 4. Results

### Long short-term memory (LSTM) selects potential phenotype-associated signals as stage 1 of the current model

To pre-select potential phenotype-associated candidates from pools of haplotype blocks in the human genome while maintaining a feasible computational burden, it is essential to implement efficient filtering techniques. These techniques should be capable of learning the representations of haplotype blocks by considering all the SNPs within each block to the phenotype. Long short-term memory (LSTM) is a recurrent neural network that is capable of learning long-range dependency and can process sequences with variable lengths. It was widely used in datasets that process “sequential” properties, such as natural language translation, before the introduction of the Transformer model (Vaswani et al., 2017). Nevertheless, LSTMs still demonstrate several advantages, especially for small datasets such as haplotype blocks, making them a potentially effective technique for the filtering stage.

In the current study, we evaluated three epistasis models (model 1, model 2, and model 3) as described by Burton et al. (2007), using chromosome 20 data from 1,000 individuals. Single nucleotide polymorphisms (SNPs) were pre-organized into haplotype blocks using *PLINK.* Detailed descriptions of the dataset simulation and haplotype block parsing methods can be found in the Methods section. Each haplotype was tested individually by LSTM (n=10) and the root of mean square error was recorded each time. Figure 3 shows the LSTM performance of chromosome 20 with 2 random haplotype blocks selected as epistasis signals of model 1, model 2 and model 3 (from top to bottom) at one record. Multiple valleys can be observed in all three plots, indicating the presence of real epistasis signals. To quantify the performance of the model across multiple runs with different haplotype block sizes, we repeated this process 10 times and recorded the ROC AUC each time. As shown in the figure, the LSTM can distinguish the epistasis signals from the background noise, achieving an area under the curve (AUC) close to 1 in all three simple epistasis models.

**Figure 3.**
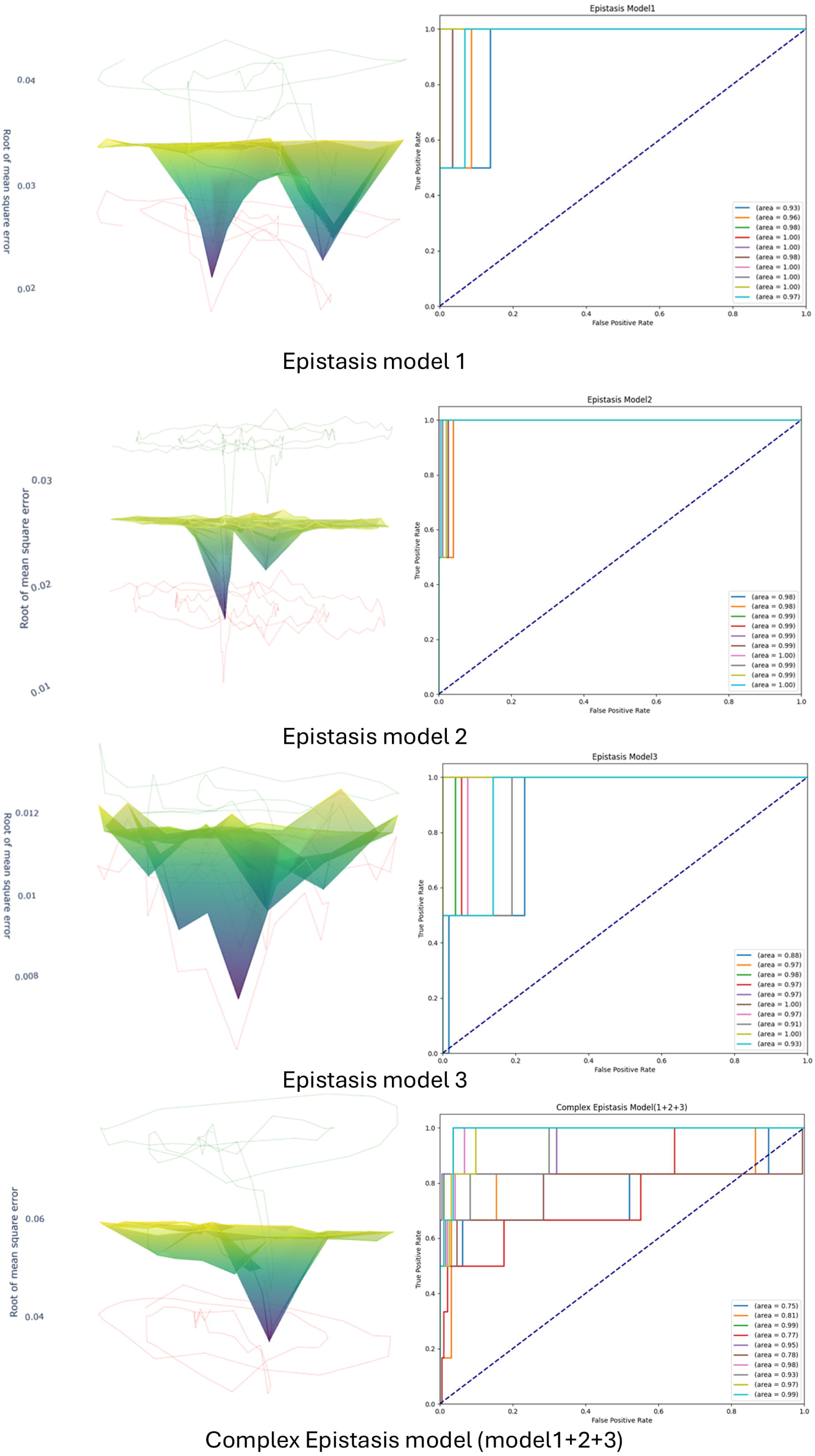
Long short-term memory (LSTM) selects potential phenotype-associated haplotype block candidates on both simple and complex epistasis models using simulated datasets. Each haplotype block was independently trained and tested by the LSTM model. The root mean square error (RMSE) of the testing datasets was plotted along with the mean and standard deviation (SD). To evaluate the model’s performance with different haplotype block lengths, the process was repeated (n=10). For each iteration, the area under the curve (AUC) of the model’s predictions was plotted against the ground truth.

Additionally, we conducted tests using the complex epistasis model to evaluate the LSTM’s capability in distinguishing signals. This complex epistasis model encapsulates the combined effects of model 1, model 2, and model 3, which feature multiple epistatic interactions from different haplotype block pairs. Employing assessment criteria similar to those used for the simple epistasis model, Figure 3 shows that the LSTM demonstrates robust performance in accurately identifying all haplotype blocks which contain complex epistasis signals. In short, in both simple and complex epistasis models, the LSTM is an effective tool for selecting potential candidates from large haplotype pools, significantly reducing the computational burden and increasing the calculation power for subsequent hierarchical transformer analysis.

### The hierarchical transformer encoder maps the haplotype block interactions with the complex epistasis model as stage 2 of the current study

After selecting the potential phenotype-associated haplotype block candidates, we applied a hierarchical transformer encoder and continued with the simulation datasets to assess its ability for epistasis signal detections. The hierarchical transformer encoder is a modified version of the hierarchical transformer that was originally proposed by Liu and Lapata (2019). The potential epistasis signals were quantified and visualised using the attention weights, which served as the main output of the current hierarchical transformer encoder. “Attention,” first introduced by Vaswani et al. (2017), is a mechanism in the transformer neural network that enables the model to dynamically weigh the importance of each element in an input sequence relative to the others. The attention weights, the main output of our current model, are computed using a scaled dot-product that quantifies this “relatedness” between pairs of haplotype blocks and SNPs (within haplotype block). Several studies (Ahmed, Aly and Liu, 2024; Graça et al., 2024) have demonstrated these attention weights that learned by the model, at least partially, can be interpreted as the epistasis interactive scores between genetic regions.

In the hierarchical transformer encoder, attention weights are initially mapped between each SNP within each haplotype block. Subsequently, a fixed-length representation of each haplotype is generated using multi-head pooling. Multi-head attention weights are then calculated between haplotypes. This updated information is incorporated into the original SNP embedding and processed through the feed-forward layer. For further details, please refer to the Methods section. Figure 4a illustrates the Root Mean Square Error (RMSE) and loss for both training and testing datasets, split in a ratio of 0.8:0.2, for a single record. The plots demonstrate a smooth training trajectory with no significant discrepancies between the training and testing datasets, indicating stable model performance and effective training without overfitting. Additionally, this process was repeated 10 times, each iteration using different pre-selected simulated signals (Figure 4b). The model consistently demonstrated a robust performance in phenotype predictions with mean RMSE lower than 0.012 for both training and testing datasets.

**Figure 4.**
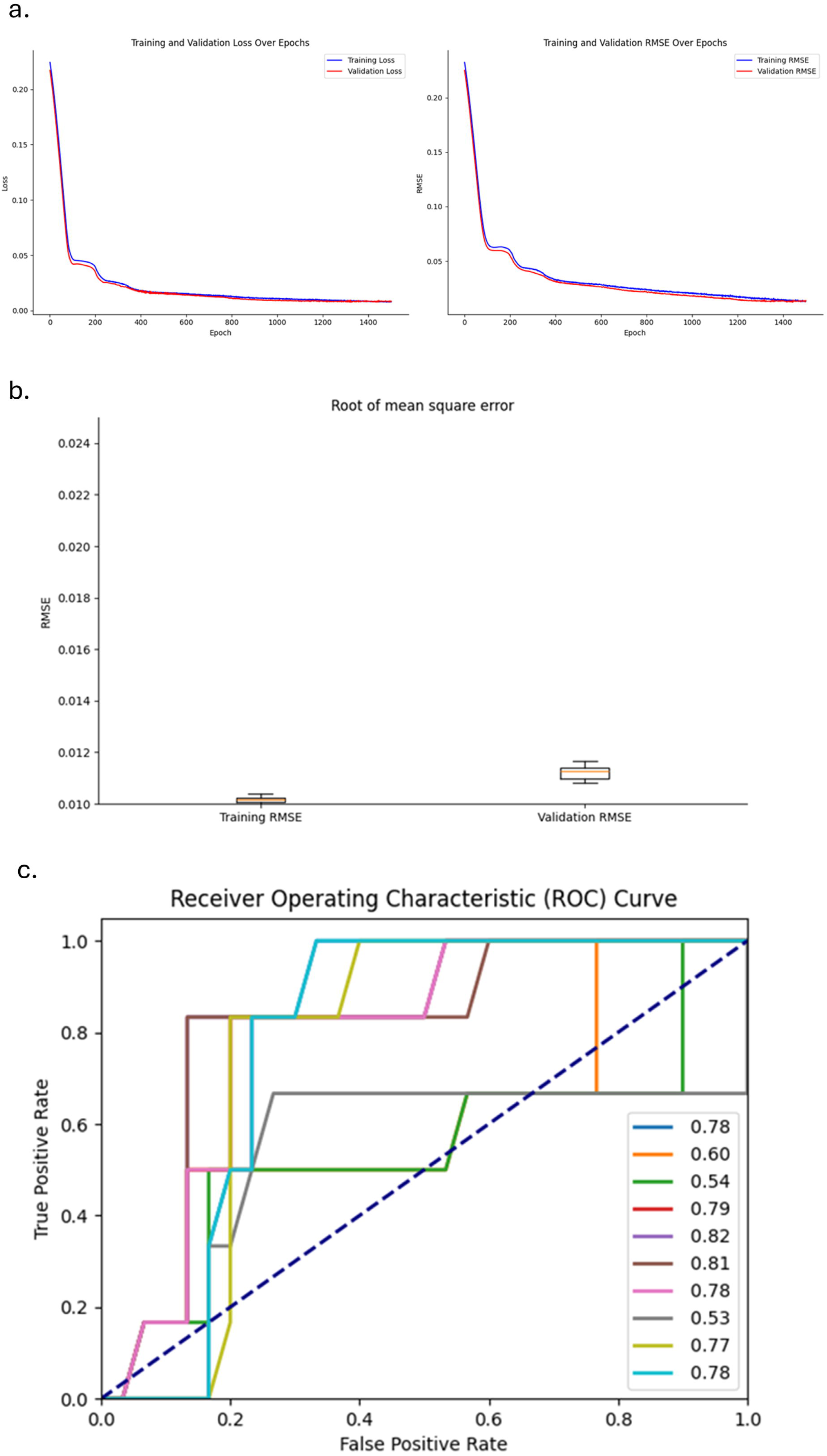
The hierarchical transformer encoder distinguishes epistasis signals in a complex epistasis model using simulated datasets. Pre-selected haplotype block candidates identified by the long short-term memory (LSTM) model were subsequently processed by the hierarchical transformer encoder. (a) Training and testing loss, and root mean square error (RMSE) for a single record. (b) Box plot showing the RMSE for training and testing datasets (n=10). (c) The area under the curve (AUC) for the complex epistasis signals predicted by the hierarchical transformer encoder, compared to the ground truth.

**Figure 5.**
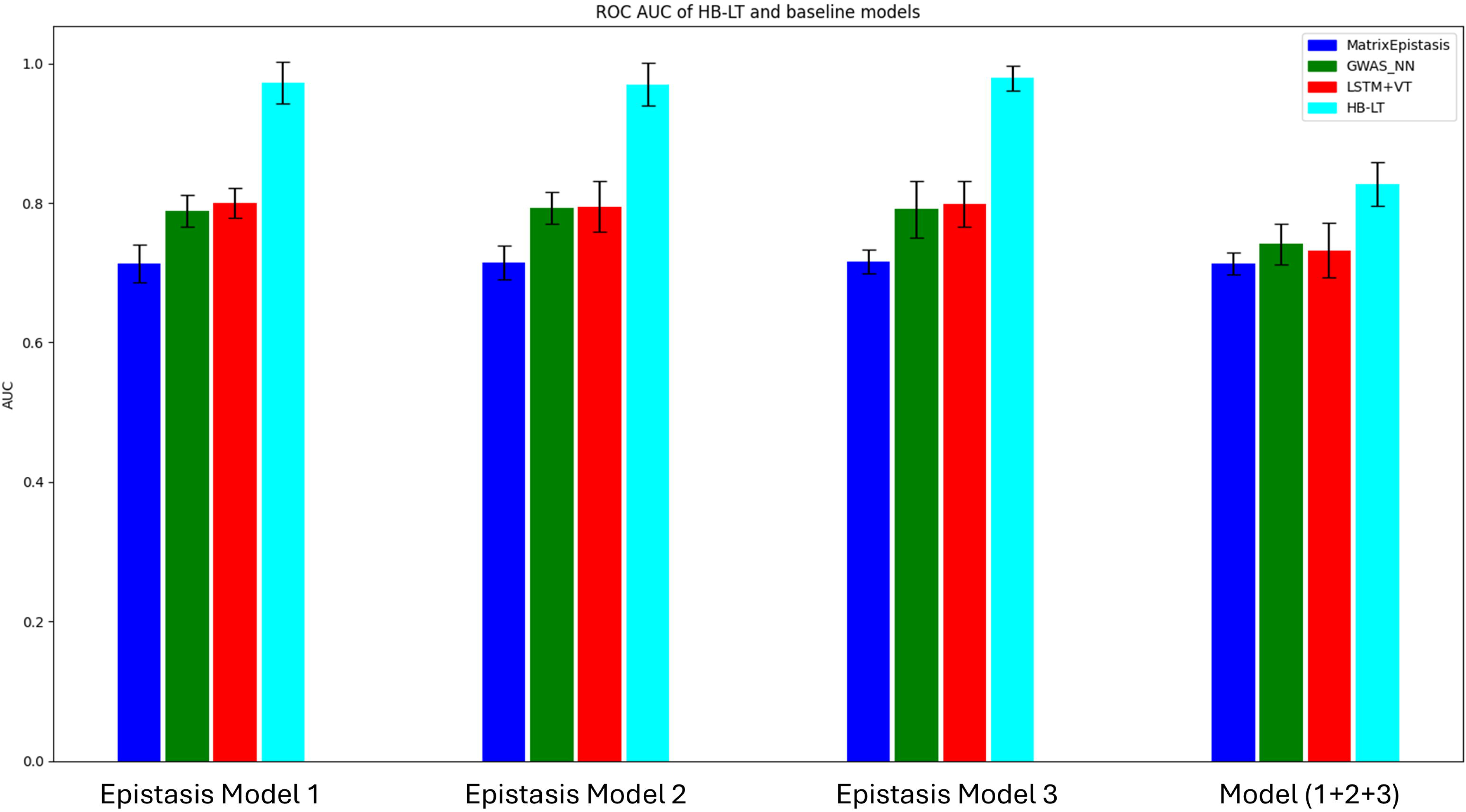
The comparison of HB-LT with state-of-art Epistasis searching methods, MatrixEpistasis (Zhu and Fang, 2017) and GWAS_NN (Cui et al., 2022) in both simple and complex epistasis models with n=10.

More importantly, to evaluate whether the simulated complex epistasis signals (model1+2+3) can be at least partially captured by the cross-haplotype attention weights, which are the primary output of our current HB-LT model, we compared these weights with the ground truth matrix. We then plotted the ROC AUC curve (n=10), as shown in Figure 4c. The model achieved an average AUC of 0.83. In short, after the LSTM pre-selected the potential phenotype-associated haplotype blocks, the hierarchical transformer encoder demonstrated reasonable performance in distinguishing potential interaction/epistasis signals between haplotype blocks.

### HB-LT outperforms baseline models for both simple and complex epistasis models

Traditional exhaustive epistasis searching methods are often facing a challenge referred to as the “curse of dimensionality”. The SNPs that are involved in the epistatic interactions might have low minor allele frequencies, however, the variants to be tested can be huge. As a result, detecting these interactions becomes challenging due to the reduced statistical power and the increased likelihood of both type 1 and type 2 errors. In the current study, we selected matrixEpistasis (Zhu and Fang, 2017) as one of the representatives for an exhaustive epistasis searching method against our current HB-LT model.

In contrast, we also tested two deep learning neural network baseline models, GWAS_NN (Cui et al., 2022) and LSTM with the vanilla transformer encoder, against our current HB-LT model. In GWAS_NN, the long sequence of SNPs was initially divided into different genetic blocks (SNPs layer). Fully connected multilayer perceptrons (MLPs) were then used to learn a fixed representation for each genetic block (*g*_1_, ..., *g_m_*). Another set of MLPs was subsequently trained to learn the epistasis between these genetic blocks. Additionally, we also tested the LSTM with the vanilla transformer encoder against our HB-LT model to assess the potential advantages of the hierarchical transformer structure compared to the vanilla single SNP transformer structure for epistasis studies.

A total of 6,156 haplotype blocks (60,501 SNPs) from chromosome 20 in the simulation datasets were analysed using three baseline models and the HB-LT model. These models were evaluated under both simple and complex epistasis conditions with recorded ROC AUC (n=10). Overall, all four models showed robust performance in identifying epistasis signals. MatrixEpistasis demonstrated a stable yet comparatively low performance, with an average of around 0.73 across both simple and complex epistasis models. This outcome likely reflects the reduced statistical power of exhaustive searching methods when handling large SNP datasets. In contrast, HB-LT exhibited the highest performance across both simple and complex models, although this advantage was less pronounced when dealing with complex models. The reason HB-LT has a larger ROC AUC than baseline models could mainly be because HB-LT employs multi-head pooling to utilise all SNPs for representing haplotype blocks in a supervised manner. This approach not only reduces dimensionality compared to considering each SNP individually but also provides more informative representations than selecting a single SNP or using unsupervised methods such as PCA.

### Interaction discovery in a diabetes glycated haemoglobin study

To inspect how trustworthy our proposed framework is in a real-world scenario, experiments on real-world cohort, UK BioBank for glycated haemoglobin (HbA1c) are performed. Glycated haemoglobin (HbA1c) is the most common biomarker used to monitor glucose control in diabetes patients (WHO, 2011), which reflects the glycemic load ∼ 3 months and traits such as hemoglobinopathies and alteration in intracellular glucose metabolism (Nathan, Turgeon and Regan, 2007). HbA1c levels are influenced by both environmental and genetic factors. Research studies (Snieder et al., 2001; Meigs et al., 2002) estimating the heritability of HbA1c in non-diabetic individuals report a range from 27% to 62%, providing strong evidence of a significant genetic component in HbA1c variability. Previous genome-wide association studies (GWAS) (Wheeler et al., 2017) have identified more than 100 genetic variants to be associated with HbA1c. In this study, we re-examined the HbA1c-associated loci in the UK Biobank cohort to explore potential novel epistasis signals, both within and across haplotype blocks. To limit the potential confounding factors in our current study, patients were pre-filtered based on 4 covariants (age of diabetes diagnosis; diabetes duration; sex and population genetic structure) in UK Biobank to select individuals with similar characteristics and eliminate the potential outliers. The clinical features of individuals from the UK Biobank used in the current study are shown in Table 1. The chosen datasets have 1277 individuals, with in total of 14 genes with 10kbp flanking regions added to both ends, 74 haplotype blocks and 1821 SNPs. The haplotype blocks were parsed based on the confidence interval method (Gabriel et al., 2002). More details regarding individual covariants pre-selection and haplotype block parsing can be referred to the Methods section.

**Table 1.** Characteristics of the included individuals of the cohorts.

|  |  |
| --- | --- |
| Clinical Features | UK Biobank (n=1277)<br>(Mean+-SD) |
| Age of diabetes diagnosis | 56.3+-3.0 |
| Duration (year) | 4.0+-2.4 |
| Diastolic blood pressure | 83.0+-5.1 |
| Systolic blood pressure | 142.0+-8.8 |
| Sex (Male: Female) | 482:795 |

### Cross-haplotype block epistasis

Unlike the simulated datasets, the interacting haplotype blocks are not known in the real datasets. One of the common approaches to validate the proposed framework’s prediction and interpretation is to find previous works that report genes and epistatic relationships on the related disease. Afterwards, the objective is to map the framework haplotype block outputs to the target genes. In the UK Biobank, using HB-LT, we observed 7 pairwise interaction candidates. All interaction candidates of the HbA1c phenotype and their corresponding attention scores were listed in the first two columns of Table 2. The threshold is set as attention scores higher than 0.1. There are no standard ways to choose the attention threshold. For the future studies, researchers can set the threshold wherever they think that fits their hypothesis.

**Table 2.**
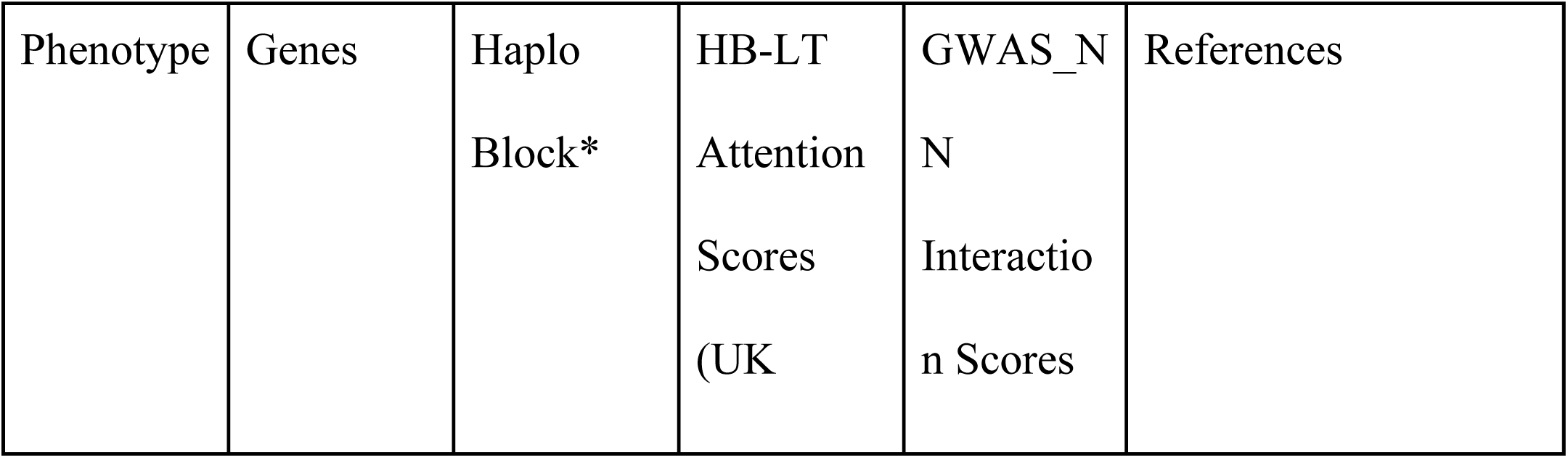
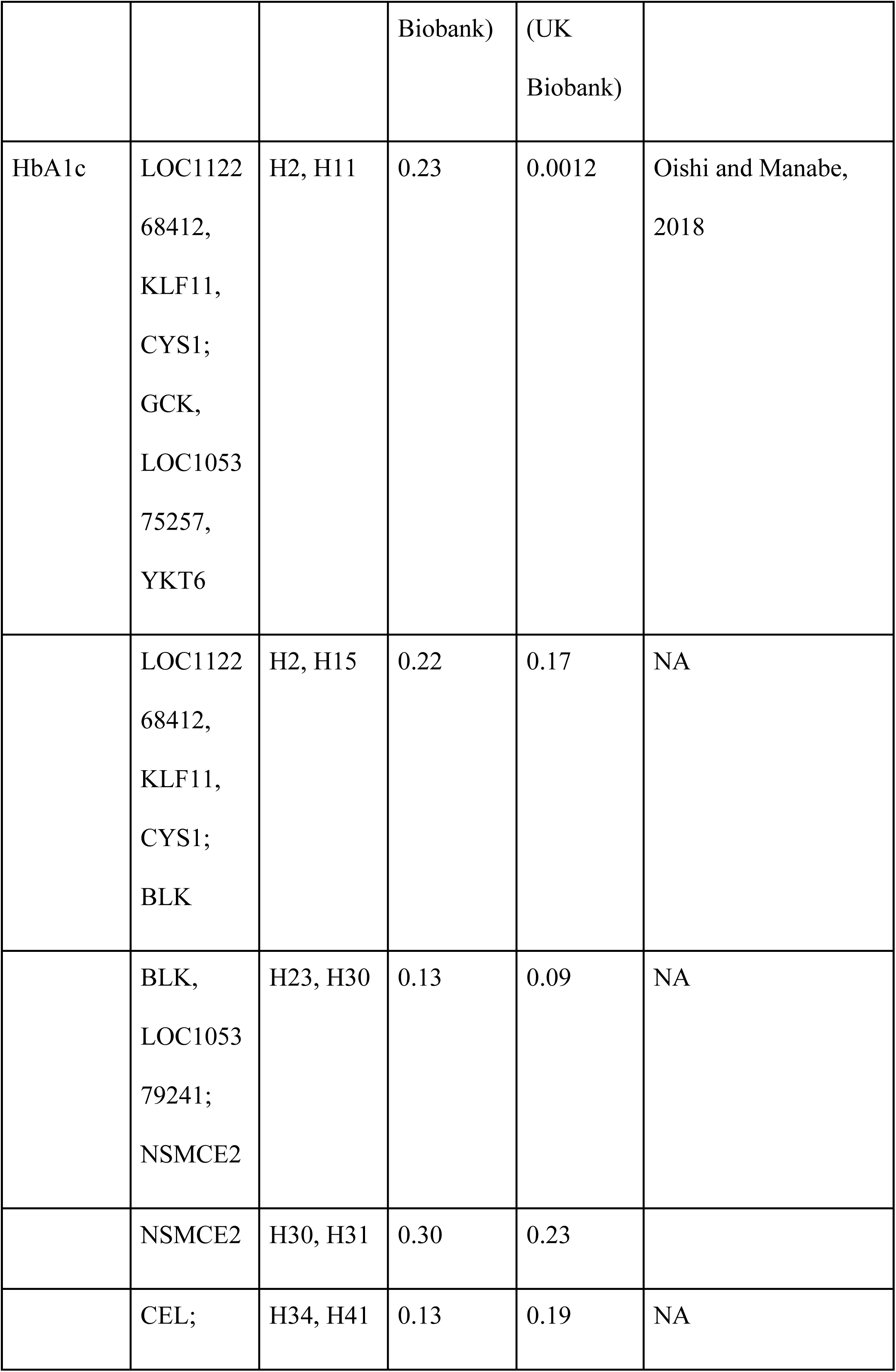
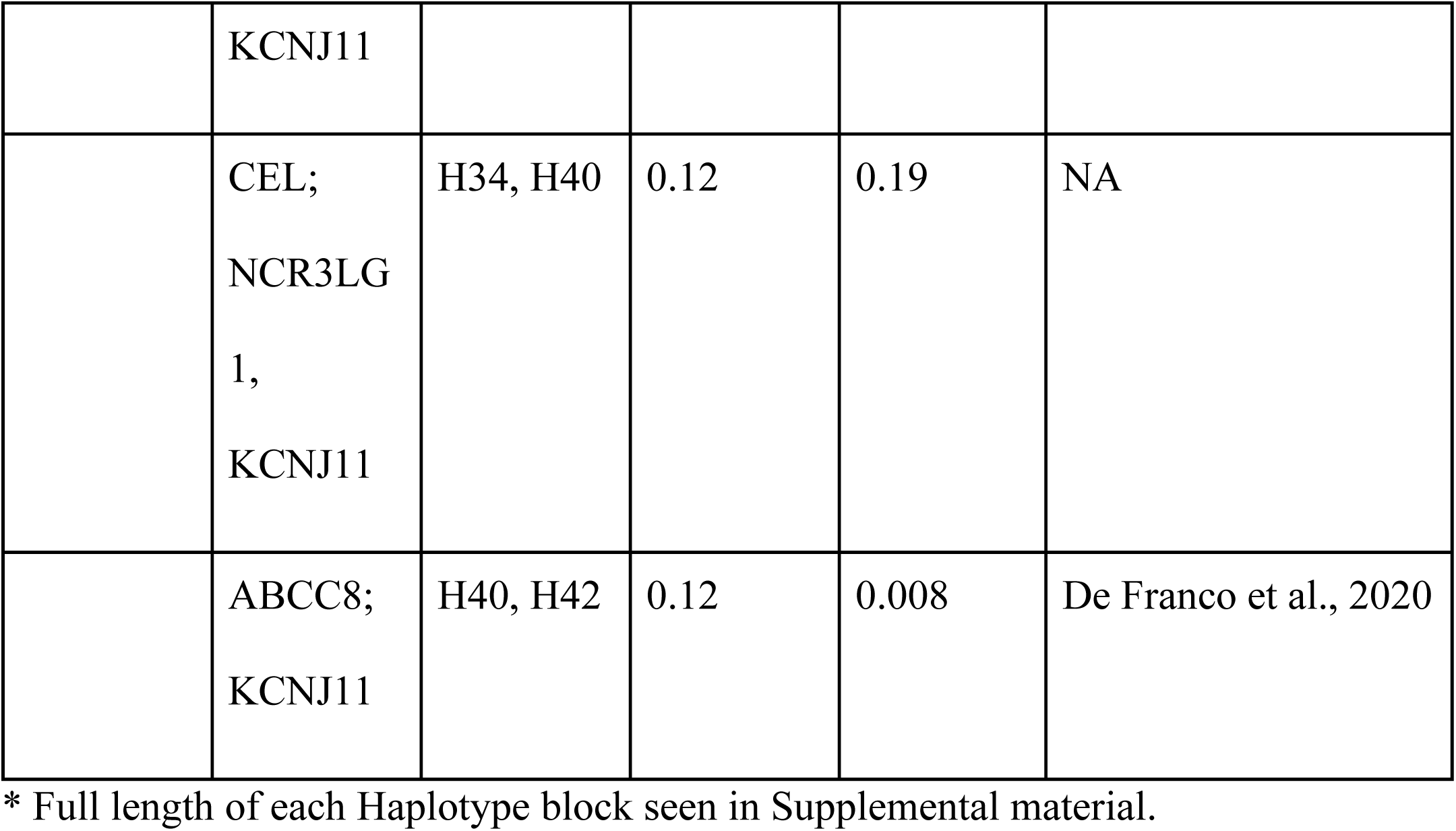
Cross-haplotype blocks attention in UK Biobank by two independent methods.

| Phenotype | Genes | Haplo<br>Block* | HB-LT<br>Attention<br>Scores<br>(UK | GWAS_N<br>N<br>Interactio<br>n Scores | References |
| --- | --- | --- | --- | --- | --- |

|  |  |  | Biobank) | (UK<br>Biobank) |  |
| --- | --- | --- | --- | --- | --- |
| HbA1c | LOC1122<br>68412,<br>KLF11,<br>CYS1;<br>GCK,<br>LOC1053<br>75257,<br>YKT6 | H2, H11 | 0.23 | 0.0012 | Oishi and Manabe,<br>2018 |
|  | LOC1122<br>68412,<br>KLF11,<br>CYS1;<br>BLK | H2, H15 | 0.22 | 0.17 | NA |
|  | BLK,<br>LOC1053<br>79241;<br>NSMCE2 | H23, H30 | 0.13 | 0.09 | NA |
|  | NSMCE2 | H30, H31 | 0.30 | 0.23 |  |
|  | CEL; | H34, H41 | 0.13 | 0.19 | NA |
|  | KCNJ11 |  |  |  |  |
|  | CEL;<br>NCR3LG<br>1,<br>KCNJ11 | H34, H40 | 0.12 | 0.19 | NA |
|  | ABCC8;<br>KCNJ11 | H40, H42 | 0.12 | 0.008 | De Franco et al., 2020 |

\* Full length of each Haplotype block seen in Supplemental material.

We then investigate whether the interaction candidates discovered by HB-LT can be detected by other methods. Similarly to the simulation dataset section, we trained GWAS_NN and recorded the interaction scores for each interaction candidate in the third column in Table 2. Not all signals detected by HB-LT can also be mapped out by GWAS_NN as significant. This could potentially highlight that HB-LT can detect novel signals which can be overlooked by other methods. Finally, We then check if any of the gene interactions have already been recorded in the previous studies, shown in the last column of Table 2.

### Within-haplotype SNP epistasis

One of the key advantages of HB-LT compared to other multi-dimensional reduction methods is that it trains each SNP individually before pooling them into a fixed haplotype representation. This approach preserves the haplotype structure during the training process, allowing us to monitor not only potential cross-haplotype interactions but also SNP interactions within each haplotype block. Indeed, it is believed that SNPs within a functional region have a higher chance to interact with each other and influence the phenotype (Ma, Clark and Keinan, 2013). We listed all the within-haplotype SNP interaction candidates, gene names and attention scores in the first three columns of Table 3. By setting the threshold > 0.05, there are 16 within haplotype block pairwise SNP interactions observed. Similarly, we compared the within-haplotype pairwise SNP interaction results obtained from HB-LT with those identified using the previously published exhaustive search method, MatrixEpistasis and recorded the *p-value* in the last column of Table 3.

**Table 3.** Within haplotype block SNP attention in UK Biobank by both HB-LT and MatrixEpistasis.

| Phenotype | Genes | Pairwise<br>SNPs | Attention<br>Scores<br>(UK<br>BioBank)<br>HB-LT | P-value (UK BioBank)<br>MatrixEpistasis |
| --- | --- | --- | --- | --- |
| HbA1c | NA | rs6756950,<br>rs7420169 | 0.11 | 0.00037 |
|  | HNF4A | rs736820,<br>rs736824 | 0.11 | 0.00046 |
|  | GCK | rs2908285,<br>rs118180640 | 0.09 | 0.00023 |
|  | YKT6,<br>GCK | rs1814253,<br>rs118180640 | 0.07 | 0.00163 |
|  | GCK, NA | rs118180640<br>,<br>rs758983 | 0.07 | 0.00091 |
|  | NEUROD<br>1, NA | rs12053195<br>,<br>rs6756950 | 0.06 | 0.052 |
|  | NA,<br>NEUROD1 | rs6756950,<br>rs12052558 | 0.05 | 0.002 |
|  | NA,<br>NEUROD1 | rs6756950,<br>rs16867467 | 0.05 | 0.0002 |

In summary, leveraging pre-selected HbA1c-associated genes from the UK Biobank, we explore the potential real-world application of HB-LT. Future studies are essential to statistically and biologically validate our current findings. Additionally, further investigation is warranted to assess the feasibility of HB-LT in hypothesis-free, large-scale genetic datasets.

## 5. Discussion

The current existing approaches for detecting interactions in genetic study face several challenges including: (i) to reduce the “curse of dimensionality”, genes are typically represented as the most important SNPs while ignoring the potential effects of neighbouring SNPs. (ii) only restrictive forms of interactions are considered. (iii) while scalable methods like PCA have been proposed to reduce the multidimensional SNP data into a one-dimensional representation, these techniques often neglect important phenotype-related information. Additionally, such methods make it difficult to analyse the internal structure of each condensed multi-SNP unit in relation to the phenotype. Indeed, there is a need for a framework that integrates genetic block representation learning with the modelling of both intra- and inter-block interactions within an end-to-end model. Here, we proposed a deep learning genetic block detection method, Haplotype Block LSTM hierarchical Transformer (HB-LT). HB-LT can hierarchically encode genetic SNP sequences. In HB-LT, each SNPs in relationship to its surrounding SNPs within each genetic block were learned by a multi-attention head. Next, a pooling method is applied to get a fixed representation of each genetic block, cross genetic block relationships via an attention method were then mapped as opposed to concatenating dimensional condensed genetic units into flat sequences and then fed into the model. This approach enables the model to dynamically learn richer structural dependencies among SNPs within each genetic block and effectively incorporate these insights into the inter-genetic block layer. In the experimental work, the results obtained from HB-LT were compared with both exhaustive searching (MatrixEpistasis) and deep learning methods (GWAS_NN). The current study shows that HB-LT exhibits a better performance for epistasis detections in both simple and complex epistasis models. Moreover, HB-LT was also tested on HbA1c in the UK Biobank to assess its application in a real-world scenario.

The current proposed deep learning framework may have many attractive features, but it also has several shortcomings. First, it should be noted, that although the potential applications of attention weights in transformer as an indicator of epistasis have been studied and demonstrated in recent years (Reyes et al., 2022; Graça et al., 2024), these weights cannot be directly interpreted as measurements of epistasis levels in genetics. While these weights can highlight regions of interest in relation to the phenotype, the focus of attention mechanisms is on capturing token dependencies and relevant patterns in the dataset to improve outcome predictions, not necessarily to isolate or quantify specific genetic interactions. Moreover, the intricate relationship between statistical and biological epistasis adds an additional layer of complexity (Moore and Williams, 2005; Ebbert, Ridge, and Kauwe, 2015). The disparity between these two models of epistasis often obscures the biological relevance and implications of statistical findings, making it challenging to draw clear, meaningful conclusions about the underlying genetic mechanisms. Indeed, we view our proposed HB-LT framework as a tool for mining and filtering large datasets. Regions of interest identified by HB-LT should be further investigated and validated through more targeted statistical methods and potential complementary biological experiments. Second, the potential covariants, such as age and population genetic structure are not directly incorporated in the current model, instead, a PCA-based pre-filtering stage was applied to minimise the confounding effects. By implementing this approach, we minimised the risk of extraneous factors contaminating the HB-LT model outputs, making the results more straightforward to interpret. However, this adjustment means that the HB-LT model will not be applied to the full dataset size, potentially leading to a loss of information. Additionally, this change may introduce new complexities for future users. The challenge of how to incorporate potential covariates into the model remains an open question that needs to be addressed. Finally, the potential of whole-genome hypothesis-free epistasis studies to significantly enhance outcome prediction has been a topic of debate and scepticism for decades. Several studies (González-Camacho et al., 2012; Mäki-Tanila and Hill, 2014 and Wei et al., 2014) have demonstrated that despite the possible biological ubiquity of epistasis, the total genetic variance of polygenic traits is likely largely to be explained by the “additive top SNPs model”.

However, other studies (Dudley and Johnson, 2009; Hu et al., 2011; Álvarez-Castro et al., 2012; Wang et al., 2012) conducted showed that the inclusion of epistatic effect networks for prediction improved prediction over the use of additive effects only. Indeed, we do not intend to propose HB-LT as a replacement for the “top SNPs approach”. This study is not aimed at comparing epistasis and non-epistasis models for outcome prediction. Instead, HB-LT serves as a complementary tool designed to uncover interactions that might otherwise be overlooked, potentially revealing novel genetic structures underlying complex disease development and leading to the discovery of new markers.

## 6. Conclusion

For the past decades, Genome-wide Association Studies (GWAS) have successfully identified thousands of risk alleles for complex diseases. Despite this, it usually failed to capture the statistical epistasis i.e. interaction between SNPs, which is acknowledged as a fundamental factor for understanding complex disease genetic pathways. Traditional epistasis searching tools often suffer from computational burden and lack of calculation power. Moreover, it has become increasingly recognized that leveraging the combined effects of SNP groups within specific genetic blocks for epistasis searching can yield greater phenotypic variance than focusing on individual SNPs alone. However, to our best knowledge, there is yet a framework that has been proposed that can incorporate this hierarchical genetic structure to form an end-to-end model for epistasis searching. Here, we proposed HB-LT, which takes advantage of the haplotype block structure existing in the genome to reduce the dimensionality of SNP features and increase statistical power. Haplotype block is not the only way to measure SNP dependencies and grouping, the impact of different methods for epistasis study should be investigated in the future.

## Funding

G.A.R. was supported by a Wellcome Trust Investigator Award (WT212625/Z/18/Z), MRC Programme grant (MR/R022259/1), Diabetes UK (BDA 16/0005485) and NIH-NIDDK (R01DK135268) project grants, a CIHR-JDRF Team grant (CIHR-IRSC TDP-186358 and JDRF 4-SRA-2023-1182-S-N), CRCHUM start-up funds, and an Innovation Canada John R. Evans Leader Award (CFI 42649). A.B. was supported by the Canada Research Chairs program.

## Duality of Interest

G.A.R. has received grant funding from, and is a consultant for, Sun Pharmaceuticals Inc. All other authors declare that there are no relationships or activities that might bias, or be perceived to bias, their work.

## Contribution Statement

The current project was designed by S.L. and S.A. under the supervision of G.A.R. and with professional support from P.H., J.T., A.B. and B.L. The current model (HB-LT) was designed by S.L. and S.A. The source code was written by S.A. and S.L. The model training and testing were performed by S.L. and S.A. The real-world cohort genetic datasets were organized, maintained and queried by R.A. The manuscript was written by S.L.

## Notes

### Competing Interest Statement

G.R. has received grant funding from, and is a consultant for, Sun Pharmaceuticals Inc. All other authors declare that there are no relationships or activities that might bias, or be perceived to bias, their work

### Author Declarations

The study used ONLY available human data from UK Biobank that were originally located at: https://www.ukbiobank.ac.uk/

### Summary of Updates

Data Availability statement revised to state that the code developed will be available at GitHub

